# SMART-HF: Structured Management Approach to Remote Treatment of Heart Failure Associated With Predictable Hemodynamic Improvements In A Community Remote Pulmonary Artery Pressure Monitoring Program

**DOI:** 10.64898/2026.04.12.26350637

**Authors:** Marc Atzenhoefer, Tamar Atzenhoefer, Hussain Boxwala, Brooke Nelson, Michelle Staudacher, Fahad Iqbal

## Abstract

**Aims:** Responses to remote pulmonary artery pressure data vary across programs. We evaluated SMART-HF, a structured pulmonary artery diastolic pressure (PAD)-guided workflow, in a community heart failure cohort.

**Methods:** We retrospectively analyzed adults with heart failure and an implanted pulmonary artery pressure sensor managed with SMART-HF. Pulmonary artery diastolic pressure (PAD) was calculated from prespecified 14-day windows at baseline, 90 days, and 6 months. Two hemodynamic management performance indices (HMPI) were prespecified: the 6-Month Delta HMPI (PAD reduction >2 mmHg from baseline) and the 90-Day Target HMPI (PAD ≤20 mmHg at 90 days). Exploratory analyses evaluated patients with baseline PAD >20 mmHg.

**Results:** Of 37 patients, 36 had paired 90-day and 29 had paired 6-month windows. Mean PAD decreased from 18.3 ± 7.0 to 16.1 ± 6.3 mmHg at 90 days and from 18.8 ± 6.8 to 15.5 ± 5.8 mmHg at 6 months (both P < 0.001). The 90-Day Target HMPI was achieved in 26/36 (72.2%) and the 6-Month Delta HMPI in 19/29 (65.5%) [95% CI 45.7-82.1]. In the exploratory subgroup (baseline PAD >20 mmHg), mean PAD changes were −2.9 ± 3.6 mmHg at 90 days (n = 19; P = 0.002) and −4.9 ± 4.9 mmHg at 6 months (n = 15; P = 0.002).

**Conclusions:** SMART-HF was associated with improved ambulatory pulmonary artery diastolic pressure control at 90 days and 6 months. Exploratory subgroup findings support further evaluation in patients with elevated baseline pulmonary artery diastolic pressure.

## 1. Introduction

The growing burden of heart failure (HF) in the United States continues to outpace multidisciplinary efforts to reduce morbidity and mortality. While 5-year survival for many malignancies now approaches 70%, those afflicted with HF are faced with persistently poor outcomes; 5-year survival after an index HF hospitalization has been reported near 25% regardless of left ventricular ejection fraction.^1,2^

Guideline-directed medical therapies (GDMT), close follow-up, and implantable pulmonary artery (PA) pressure monitoring have been established as effective tools to reduce congestion-related HF events. However, real-world adoption and execution of these proven interventions remain heterogeneous, with management differing materially between providers and systems. Even when real-time hemodynamic data are available, treatment decisions may still rely on delayed clinical surrogates (symptoms, weight changes, and examination findings), delaying recognition and treatment of congestion.

Evidence linking ambulatory PA pressures to outcomes is no longer theoretical. Large pooled analyses have demonstrated that ambulatory PA pressures and their change over time independently predict subsequent mortality.^4^ Complementary evidence from real-world datasets suggests that under ad hoc management, a minority of patients with elevated baseline PAD achieve PAD thresholds associated with improved survival^4,5^—highlighting an implementation gap that may be addressable with standardized, reproducible workflows.

We propose SMART-HF (Structured Management Approach to Remote Treatment of Heart Failure), a PAD-centric, algorithm-driven workflow designed to reduce variability in response to remote PA pressure data and systematically drive PAD toward physiologically normal targets.

The primary aim of the present study was to evaluate the hemodynamic effectiveness of SMART-HF by quantifying change in PAD and PAD target attainment using prespecified 14-day averaged windows.

## 2. Methods

### 2.1 Study design and setting

Single-center retrospective cohort analysis of adults with heart failure implanted with a remote pulmonary artery pressure sensor and managed exclusively using the SMART-HF workflow at Froedtert Menomonee Falls Hospital (Menomonee Falls, Wisconsin, USA). Implant dates in the analyzed cohort ranged from 2025-02-11 to 2025-08-28.

Ethics/oversight: Institutional review board reviewed; informed consent waived.

### 2.2 Participants

Eligibility followed current United States Food and Drug Administration indications for the CardioMEMS HF System. Under original labeling, patients with New York Heart Association (NYHA) class III heart failure and a heart failure hospitalization within the preceding 12 months were eligible. Under expanded indications, patients with NYHA class II heart failure without a recent heart failure hospitalization were eligible if N-terminal pro-B-type natriuretic peptide (NT-proBNP) met body mass index-indexed thresholds stratified by left ventricular ejection fraction category. All adults (≥18 years) with an implanted remote pulmonary artery pressure sensor at Froedtert Menomonee Falls Hospital who were managed exclusively via SMART-HF were included. No additional exclusion criteria were applied beyond standard contraindications to pulmonary artery pressure sensor implantation.

Sex was ascertained from the electronic health record and reported as a baseline characteristic (Table 1). Both male and female patients were included without restriction. The modest sample size (n = 37) precluded pre-specified sex-stratified analyses of hemodynamic outcomes; however, sex was examined as a descriptive covariate.

**Table 1.**
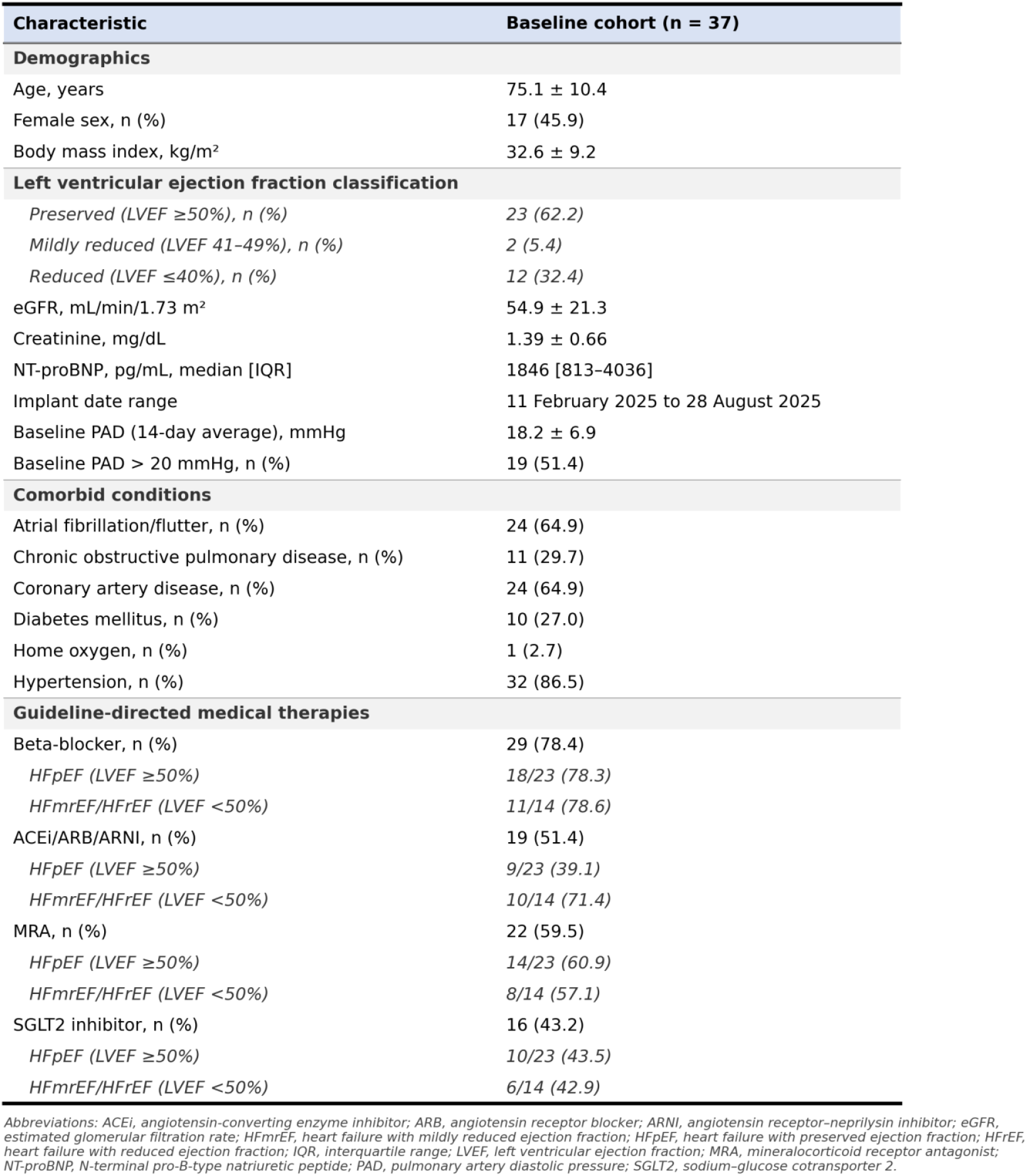
Baseline characteristics (baseline pulmonary artery diastolic pressure window cohort)

**Table 2.**
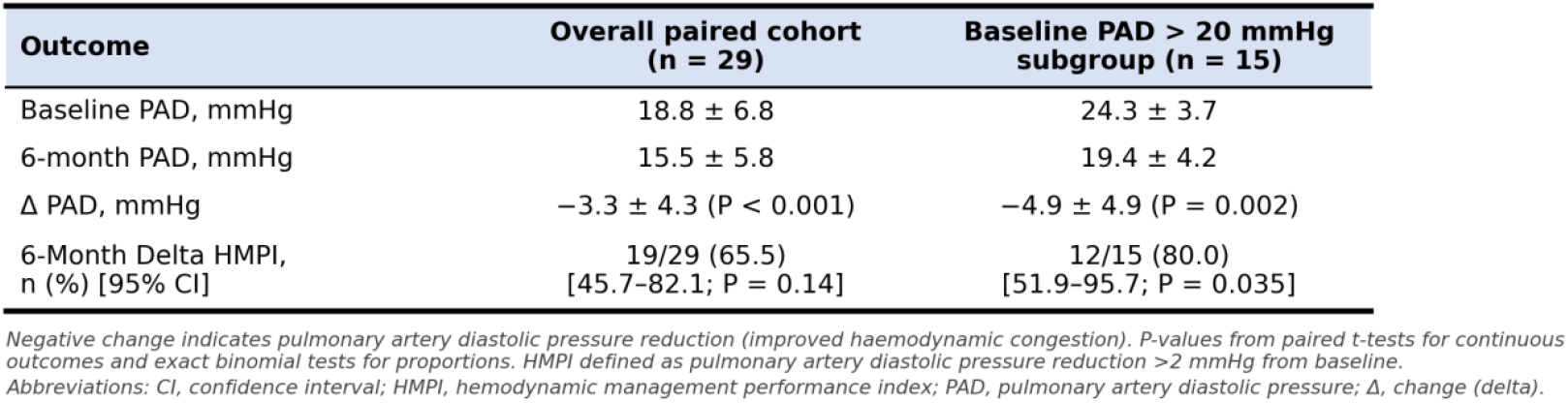
Hemodynamic outcomes (paired 6-month analysis)

### 2.3 SMART-HF workflow

Operationalized review of the entire cohort was performed in accordance with previously published methodology. ^3^ Remote pulmonary artery pressures were reviewed by advanced practice providers every Monday, Wednesday, and Friday. Patient-specific pulmonary artery diastolic pressure targets were set at time of sensor implantation. For patients with right heart catheterization PAD values of 25 mmHg or lower, 15 mmHg was designated as goal pressure. For individuals with significant congestion evidenced by invasive PAD pressures of >25 mmHg, measured PAD value minus 10 mmHg derived the initial target pressure which was subsequently refined during the early post-implant period (Days 5-7) with ideal goal for most patients being 15 mmHg.

Pharmacologic therapy was augmented according to prespecified triggers and thresholds after three serial pulmonary artery diastolic pressure transmissions met criteria relative to the patient’s target. Triggers and actions were as follows: pulmonary artery diastolic pressure ≥5 mmHg below target prompted stopping metolazone (if used) and/or decreasing loop diuretic frequency; pulmonary artery diastolic pressure 3-5 mmHg above target prompted increasing loop diuretic frequency to twice daily for two days and/or (if on furosemide) transitioning to an equivalent dose of torsemide or bumetanide; pulmonary artery diastolic pressure ≥6 mmHg above target prompted adding metolazone (renal-adjusted dose) and/or doubling loop diuretic frequency for three days and/or switching to torsemide or bumetanide with an approximately 25% dose increase. Additional safety rails, maximum daily dosing, and laboratory monitoring are summarized in Figure 1.

**Figure 1.**
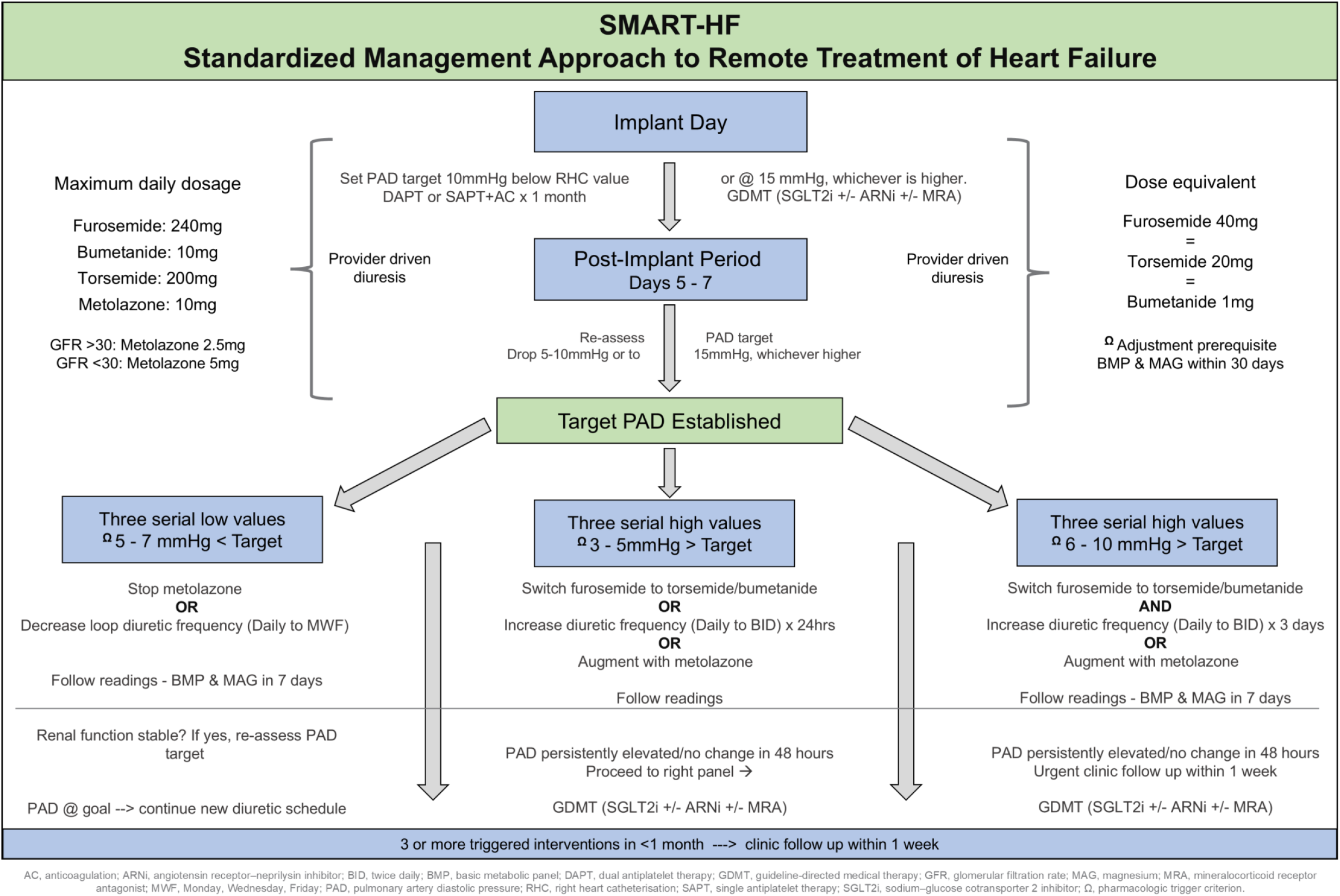
SMART-HF structured pulmonary artery diastolic pressure–guided workflow.

### 2.4 Hemodynamic data and pulmonary artery diastolic pressure windows

Pulmonary artery diastolic pressure was evaluated according to previously described 14-day windows wherein all available transmitted values within prespecified window were averaged. ^4^ Baseline pulmonary artery diastolic pressure was defined as the mean during Days 1-14 post-implant. The 90-day pulmonary artery diastolic pressure window was defined as Days 76-89, and the 6-month window was defined as Days 166-179. Averaged values were included in analysis if they contained ≥5 transmissions at baseline and ≥4 transmissions in the 90-day or 6-month window as defined in prior publications. ^4,5^

### 2.5 Outcomes

The primary outcome was change in pulmonary artery diastolic pressure from baseline to approximately 6 months using paired 14-day windows. Secondary hemodynamic outcomes included change in pulmonary artery diastolic pressure at 90 days. Two hemodynamic management performance indices (HMPI) were prespecified. The 90-Day Target HMPI was defined as the proportion of patients achieving pulmonary artery diastolic pressure ≤20 mmHg at 90 days; among patients with elevated baseline pressures (>20 mmHg), this transition has been independently associated with lower all-cause mortality.^5^ The 6-Month Delta HMPI was defined as the proportion achieving a pulmonary artery diastolic pressure reduction >2 mmHg from baseline to 6 months, a threshold associated with a 14.7% decrease in mortality risk.^4^ Exploratory subgroup analyses evaluated the same outcomes and HMPI in patients with baseline pulmonary artery diastolic pressure >20 mmHg.

### 2.6 Statistical analysis

Continuous variables are reported as mean ± standard deviation or median [interquartile range], as appropriate. Paired changes in pulmonary artery diastolic pressure were assessed using paired t-tests. Proportions are reported with exact binomial 95% confidence intervals (CI) with corresponding two-sided P-values from the exact binomial test. Statistical software: R version 4.5.3 (R Foundation for Statistical Computing, Vienna, Austria).

## 3. Results

### 3.1 Cohort and analyzable windows

Thirty-seven patients met the baseline transmission threshold (≥5 readings in Days 1-14). Thirty-six had paired baseline and 90-day analyzable windows (≥4 readings in Days 76-89), and 29 had paired baseline and 6-month analyzable windows (≥4 readings in Days 166-179). Eight patients lacked an analyzable 6-month window because no transmissions were yet available as recent date of implantation precluded any possibility of 6-month window at the time of data analysis and manuscript drafting. One patient was excluded from the 90-day analysis because only three transmissions were available in the window.

### 3.2 Baseline characteristics

Baseline characteristics are shown in Table 1.

### 3.3 Primary hemodynamic outcome

In the paired 6-month cohort (n=29), mean baseline PAD was 18.8 ± 6.8 mmHg and mean 6-month PAD was 15.5 ± 5.8 mmHg, corresponding to a mean change of −3.3 ± 4.3 mmHg (95% CI −4.9 to −1.7; P< 0.001).

### 3.4 Secondary hemodynamic outcomes

In the paired 90-day cohort (n= 36), mean pulmonary artery diastolic pressure decreased from 18.3 ± 7.0 to 16.1 ± 6.3 mmHg (mean change −2.1 ± 3.3 mmHg; 95% CI −3.2 to −1.0; P< 0.001). The 90-Day Target HMPI was met in 26/36 (72.2%) [95% CI 54.8-85.8; P= 0.011], and a pressure reduction >2 mmHg was observed in 15/36 (41.7%) [95% CI 25.5-59.2; P= 0.41]. In the paired 6-month cohort (n= 29), the 6-Month Delta HMPI was met in 19/29 (65.5%) [95% CI 45.7-82.1; P= 0.14].

### 3.5 Exploratory subgroup analysis: baseline pulmonary artery diastolic pressure >20 mmHg

Nineteen of 37 patients (51.4%) had baseline pulmonary artery diastolic pressure >20 mmHg and comprised the exploratory subgroup. All 19 had paired 90-day windows; 15 had paired 6-month windows. At 90 days (n= 19), mean pulmonary artery diastolic pressure decreased from 23.6 ± 3.5 to 20.7 ± 4.2 mmHg (mean change −2.9 ± 3.6 mmHg; 95% CI −4.7 to −1.2; P= 0.002). The 90-Day Target HMPI was met in 9/19 (47.4%) [95% CI 24.4-71.1], and a pressure reduction >2 mmHg was observed in 9/19 (47.4%) [95% CI 24.4-71.1]; these proportions were coincidentally identical but did not represent entirely the same patients. At 6 months (n = 15), mean pulmonary artery diastolic pressure decreased from 24.3 ± 3.7 to 19.4 ± 4.2 mmHg (mean change −4.9 ± 4.9 mmHg; 95% CI −7.6 to −2.2; P= 0.002). The 6-Month Delta HMPI was met in 12/15 (80.0%) [95% CI 51.9-95.7; P= 0.035].

## 4. Discussion

In this community program, SMART-HF—a structured, algorithm-driven workflow for remote pulmonary artery pressure management—produced statistically significant reductions in ambulatory pulmonary artery diastolic pressure at both 90 days and 6 months. Unlike ad hoc approaches in which clinician responses to hemodynamic data vary by provider, day, and institution, SMART-HF standardizes the entire chain of action: target setting, review cadence, threshold-based pharmacologic triggers, and safety monitoring. The result is a safe reproducible system that can be taught, audited, and scaled—addressing the well-documented implementation gap between the availability of remote hemodynamic data and its translation into consistent, evidence-aligned therapy.^3^

Initial description of the operational framework underpinning SMART-HF was in a 2025 publication. For the first time, a comprehensive unambiguous approach to remote pulmonary artery pressure management had been provided. Every step of the process was detailed starting with target pressure selection at implantation, the algorithm compelled aggressive early post-implant pressure reduction, further mandating early and frequent reassessment of goal pressures to a target of no more than 15 mmHg. Explicit guidance continued all the way down to patient specific individualization of hemodynamic triggers, diuretic strategies, all while maintaining safety through prescribed guardrails.^3^ The present analysis extends this foundational work by demonstrating that SMART-HF achieves hemodynamic performance benchmarks that have subsequently been independently linked to mortality reduction.^4,5^

Prior publications shared, in the broadest of terms, management guidance for clinicians to reduce filling pressures using diuretics and vasodilators without specifying how, when, or by how much.^10^-^12^ The distinction is beyond semantic - SMART-HF is a detailed field guide for the effective utilization of precision device guided heart failure therapy optimized without regard to resource availability or practice setting.

Two independent large-scale analyses have now established that ambulatory pulmonary artery diastolic pressure trajectories predict all-cause mortality.^4,5^ A pooled analysis of pivotal trial cohorts demonstrated that a pressure reduction >2 mmHg from baseline to 6 months conferred a 14.7% decrease in 2-year mortality, whereas an increase >2 mmHg predicted a 26.7% increase.^4^ Separately, a national Medicare cohort analysis showed that among patients with elevated baseline pressures (>20 mmHg), only 24% transitioned to acceptable levels (≤20 mmHg) within 90 days under prevailing management; yet those who did experienced significantly improved survival (hazard ratio [HR] 0.72; 95% CI 0.64-0.81; P< 0.001).^5^ These findings define the hemodynamic management performance indices (HMPI) against which any structured workflow must now be measured.

Measured against these benchmarks, SMART-HF achieved materially superior hemodynamic performance. The 90-Day Target HMPI was met in 26/36 (72.2%) of the overall cohort and in 9/19 (47.4%) of patients with baseline pressures >20 mmHg—approximately double the 24% transition rate observed in the Medicare cohort under ad hoc management.^5^ The 6-Month Delta HMPI was met in 19/29 (65.5%) overall and 12/15 (80.0%) of the elevated-baseline subgroup (P = 0.035), with a mean 6-month pressure reduction of −3.3 mmHg across the cohort—nearly double the −1.8 mmHg observed in the Medicare cohort and exceeding the −2.0 mmHg reported in pooled pivotal trial populations.^4,5^ Extrapolation of these mortality associations to the present cohort suggests that SMART-HF would confer mortality benefit to a substantially larger proportion of patients than has been achieved under any previously described management approach. The magnitude of pulmonary artery diastolic pressure reduction was greater at 6 months than at 90 days and was greatest among patients with baseline pressures >20 mmHg, suggesting that our structured approach drives continued hemodynamic improvement beyond the early post-implant period. Notably, this cohort — characterized by advanced age and multiple comorbid conditions — represents the patient population frequently cited as frail and difficult to manage with a hemodynamic guided treatment strategy through remote pulmonary artery pressure monitoring.

No significant change in the number or dose of guideline-directed medical therapies prescribed from baseline compared to 90 days or 6-month end points was observed in our cohort. Loop diuretics are essential for relief of congestive symptoms in heart failure, but unlike disease-modifying therapies, their long-term effect on survival and major heart-failure events is uncertain.^6^ The prevailing view that diuretics have no impact on mortality is based on small, dated placebo-controlled trials that predate contemporary guideline-directed medical therapies. Whether effective diuretic therapy achieving true cardiac decongestion influences disease progression and survival remains unresolved.^6^ Early signals that challenge any prognostic benefit came from a stable-heart-failure withdrawal study and later observational analyses in which higher diuretic requirements were associated with worse survival. Importantly, those analyses were unable to separate treatment effect from underlying disease severity.^7,8^ A larger, more recent trial demonstrated no significant difference in mortality or hospitalization between torsemide and furosemide after heart-failure hospitalization, reinforcing the prevailing view that loop diuretics have no impact on morbidity or mortality.^9^ Critically, none of these studies had the benefit of continuous hemodynamic monitoring afforded by an implanted pulmonary artery pressure sensor. In contrast, landmark trials of hemodynamic-guided management showed reduced heart-failure hospitalizations, and medication analyses suggested that targeted changes in diuretics and vasodilators were central to that benefit.^10^-^12^

Taken together, these observations reframe the diuretic-mortality question. Prior studies asked whether a specific loop diuretic improves survival and found no signal—but none had the ability to measure whether the diuretic actually achieved its hemodynamic objective. SMART-HF demonstrates that when diuretic therapy is titrated with precision against a continuously measured hemodynamic target, pulmonary artery pressures can be reduced reliably and sustainably. The mortality data now available indicate that these pressure reductions—regardless of the specific pharmacologic agent used to achieve them—are independently associated with improved survival.^4,5^ The critical variable may not be which diuretic is prescribed, but whether it is dosed and adjusted with sufficient precision to achieve and maintain hemodynamic targets that matter.

Our structured approach to drive down pulmonary artery pressures resulted in no serious adverse event. There were no hospitalizations or complications directly related to SMART-HF. Mild electrolyte derangements, specifically hypokalemia, hyperkalemia, and hypomagnesemia, were encountered during the observation period. In total there were seven patients with electrolyte derangements that mandated additional management. Hyperkalemia related to initiation of spironolactone was seen in two patients with resolution once agent was discontinued. The remaining five occurrences of hypokalemia and hypomagnesemia were attributed to diuretic induced renal losses. A single patient with hypokalemia from loop diuretics had spironolactone added to correct for hypokalemia. There were two patients with hypokalemia and concurrent hypomagnesemia attributed to metolazone administration. Oral supplementation of potassium and magnesium was sufficient to address these cases. The remaining two patients had either isolated hypokalemia or hypomagnesemia unrelated to metolazone administration similarly corrected by oral supplementation.

Labile renal indices which met Kidney Disease Improving Global Outcomes (KDIGO) criteria for acute kidney injury (AKI) were observed in eight patients during the observation period. All eight occurrences were during active decongestion; all eight patients had pulmonary artery diastolic pressures >22 mmHg when creatinine elevations occurred. Underlying etiology of AKI in five of eight patients was determined to be from occult paroxysmal atrial fibrillation (AF). Transmitted pressure waveforms were suspicious for atrial arrhythmia and subsequently confirmed by ambulatory electrocardiographic monitoring. Electrophysiology referral for rhythm control strategy was successful in all five of these patients with oral antiarrhythmic therapy. With restoration of normal sinus rhythm, pulmonary artery diastolic pressures fell precipitously without additional intervention or recurrent renal dysfunction. The remaining three patients were ultimately referred to nephrology with presumed diagnoses. Consulting experts confirmed diabetic chronic kidney disease (CKD) in two patients for which continued reduction of pulmonary artery diastolic pressures did not result in further elevation of serum creatinine. The third patient was diagnosed with acute tubular necrosis related to hypotension attributed to combination GDMT. After discontinuation of their sodium-glucose cotransporter 2 (SGLT2) inhibitor and decrease in dose of valsartan-sacubitril, serum creatinine returned to baseline.

While cross-study comparisons warrant caution given differences in cohort size and follow-up, the direction and magnitude of these findings are unambiguous. Under SMART-HF, every prespecified hemodynamic management performance index exceeded the benchmarks observed in both a Medicare real-world cohort and pooled pivotal trial populations.^4,5^ The question is no longer whether structured hemodynamic management improves pressure trajectories relative to ad hoc approaches—it does. What remains is to determine whether the hemodynamic gains demonstrated here translate into the survival differences that the underlying mortality associations would predict.^4,5^ Until adequately powered comparative trials are completed, SMART-HF provides a fully operationalized, reproducible framework that can serve as the standard against which future hemodynamic management strategies are evaluated.

### 4.1 Limitations

This single-center study is modest in size (n = 37). Incomplete paired follow-up was present for one patient at 90 days and eight patients at 6 months, though the reasons for each are known. The single patient missing from the 90 day comparison did not meet the 4 or more measurement transmission requirement during the 14 day follow up window per previously published criteria.^4^ The eight patients without 6-month paired data had been implanted fewer than six months before data extraction. Although this missingness is non-informative, it has the potential to introduce bias. No contemporaneous control group was studied in parallel although prior large-scale randomized trials have consistently demonstrated the efficacy of hemodynamic-guided management versus usual care. Nevertheless, our results should be interpreted for feasibility and safety rather than causal proof of superiority of SMART-HF over other management approaches.

## 5. Conclusions

SMART-HF, a structured pulmonary artery diastolic pressure-guided workflow first described in detail in 2025,^3^ produced statistically significant pressure reductions and exceeded independently validated hemodynamic management performance indices at both 90 days and 6 months in a community heart failure cohort characterized by advanced age and multimorbidity. Performance against these mortality-linked benchmarks was materially superior to that observed in both a national Medicare cohort and pooled pivotal trial populations under ad hoc management.^4,5^ These findings support the adoption of structured, algorithmic hemodynamic management as the operational standard for remote pulmonary artery pressure monitoring programs. Adequately powered comparative trials are warranted to confirm whether the hemodynamic advantages demonstrated here translate into the survival benefit that current mortality data would predict.

## Data availability

The de-identified data underlying this article will be shared on reasonable request to the corresponding author, subject to institutional and ethical approvals.

## Ethics

The study was reviewed by the local institutional review board, which waived informed consent for this retrospective analysis conducted in accordance with the Declaration of Helsinki.

